# Exploring Perceptions, Challenges, and Opportunities of UDS Health Professionals’ Students and Faculty Regarding Community-Based Education Programs for Interprofessional Education: A study protocol

**DOI:** 10.1101/2025.08.01.25332714

**Authors:** Maxwell Ateni Assibi, Bruce Abugri Ayabilla, Shamsu-Deen Ziblim, Patience Afua Adwaapa Karikari, Victor Mogre

## Abstract

**Background:** Interprofessional Education (IPE) is essential in building Interprofessional collaboration and improving patient outcomes within the healthcare industry. Despite its increasing international adoption, little evidence exists on students’ and faculty especially those within low-resource settings, perceptions and understanding of IPE.

The University for Development Studies (UDS) of Ghana, through its community-based education activities such as the Community-Based Education and Service (COBES) and the Third Trimester Field Practical Programme (TTFPP), gives the means for experiential, inter-professional learning.

This study aims to clarify the ways UDS health professional students and faculty view, experience, and navigate IPE in these courses with an emphasis on significant challenges and opportunities within a limited resource setting.

**Methods:** A qualitative approach will be utilized. In-depth interviews will be conducted among purposively selected students and faculty involved in community-based education and services (COBES) and Third Trimester Field Practical Programme (TTFPP). The data will be analyzed thematically to indicate reiterating perceptions, challenges and opportunities.

**Anticipated findings:** Both health professionals’ students and faculty will be expected to enjoy the programs for fostering collaboration, communication, and team learning. However, problems will likely occur with the lack of regular schedules, minimal resources, faculty readiness, and professional hierarchies reported in literature. Great opportunities may exist in systematic Interprofessional projects, reflective sessions, collaboration via online platforms, and support structures by faculty.

**Conclusion:** The study hopes to inform the strengthening of community-based programs to enhance Interprofessional learning. Enhanced strengthening of the programs is potentially able to prepare a health workforce that can provide coordinated care in different settings.

## Introduction

Interprofessional Education (IPE) is a learning approach that is collaborative in nature and the purpose is to facilitate health professionals’ collaboration towards better delivery of healthcare. The World Health Organization, (2010) defined interprofessional education as “learning about, from, and with each other to enable effective collaboration and improve health outcomes” (WHO. 2010). This definition identifies the essential role of interprofessional education in the development of interprofessional competencies towards patient-centered and holistic care (Suwanchatchai et al., 2024). Research has demonstrated that interprofessional education enhances collaborative potency, better patient outcomes, and fosters a healthier workforce through proper communication, collaboration, and decision-making among health professionals (Julie et al., 2016; Thistlethwaite et al., 2014). Nevertheless, IPE implementation is hindered by daunting obstacles that include resistance to interprofessional education, logistical constraints, and poorly prepared faculty (Sunguya et al., 2014; Hammick et al., 2007; Thistlethwaite et al., 2014).

The benefits of interprofessional collaboration are far-reaching and presented in detail, encompassing enhanced patient-centered care, optimum provision of health services, and enhancing the working-together culture in practice (Green BN & Johnson CD., 2015). The WHO (2010) encourages IPE as one of the significant solutions for bridging gaps in healthcare as well as enhancing the effectiveness of health systems, particularly in resource-poor settings. However, since more than mere curriculum reform is required to facilitate effective implementation of IPE, it should be accompanied by change of culture in institutions, adequate resources, and a commitment to breaking down entrenched professional silos (Hammick et al., 2007; Thistlethwaite et al., 2014).

Community-Based Education (CBE) is a teaching methodology that integrates academic learning with practical experience in community settings. CBE emphasizes pragmatic, experiential learning in which learners engage in direct contact with communities to address real health and development issues (Julie et al., 2016; Suwanchatchai et al., 2024). In Ghana, the University for Development Studies (UDS) is the pace setter in community-based education and is presently operating two principal programs: the Third Trimester Field Practical Programme (TTFPP) and the Community-Based Education and Service (COBES). The programs are designed to combine professional training and social responsibility, encouraging students to engage in interdisciplinary, community-based practice (Mohammed & Yirbekyaa, 2018; Amalba et al., 2020).

The Community-Based Education (CBE) approach promotes experiential learning through actual health setting immersion of learners. Through community-based learning exercises, Community-based education equips health professional students with skills to address healthcare challenge and fosters interprofessional collaboration (Julie et al., 2016). Through Community-Based Interprofessional Education (CBIPE), students from different health professions work together on health projects for their communities which enhances their skills in social determinants of health and social responsibility according to the IPE norms (Suwanchatchai et al., 2024). Suwanchatchai et al., (2024) found that the students engaged in CBIPE showed improved problem-solving, flexibility and greater appreciation of interprofessional collaboration in health care delivery. For effective implementation of IPE, however, institutional support is considered critical and requires appropriate resources and community engagement (Suwanchatchai et al., 2024).

The Community-Based Education and Service (COBES) model illustrates how interprofessional education (IPE) integrates with health profession education. COBES immerses students into community settings for an interdisciplinary level learning experience that reinforces interprofessional collaboration at the core (Suwanchatchai et al., 2024), placing patient-centered care and social accountability at the forefront. This type of learning allows students to acquire collaborative practice skills and develop an understanding of the roles and contributions of different health professionals in addressing community needs (Hosny et al., 2013). Furthermore, Suwanchatchai et al. (2024) again demonstrated that COBES improves the collaborative leadership and conflict resolution skills among the professions that allow them to integrate easily into various healthcare settings.

In addition to the COBES model, the Third Trimester Field Practical Programme (TTFPP) another community-based learning program for students across diverse disciplines within the University for Development Studies, including health professionals. The programmes is inbuilt in the legislation that constitutes the university (PNDC law 279, 1992). The TTFPP is an experience that encourages group work at the community level and experiential learning, fostering interdisciplinary collaboration and social accountability (Mohammed et al., 2018). It brings students from diverse academic backgrounds to engage in community development tasks, where they learn from, and with one another to address community needs in areas such as health, education and social amenities (Mohammed et al., 2018). The TTFPP fosters problem-solving, adaptability, and comprehension of the social determinants of health. Although it shares similar objectives with COBES, its interprofessional nature presents a worthy opportunity for optimizing interprofessional education.

Despite the existing literature on IPE, there is insufficient evidence linking these community-based education programs (COBES and TTFPP) to IPE to develop robust educational models that address global health priorities and strengthen health systems through interprofessional collaboration. The sustainability of the community-based education programs, however, require ongoing research, policy, and institutional investment in interprofessional training.

### Problem Statement

Interprofessional Education (IPE) is an important way of getting health professionals to work together effectively. Community-based education and services (COBES) immerses medical students into community experiential learning environment. The Third Trimester Field Practical programme (TTFPP) on the other hand, places students of various disciplines including health professionals students into communities during the first year and second years of their study period. Despite the many years of practicing these community-based education programs by the University for Development Studies, there is lack of studies creating awareness on these programs for promoting interprofessional education. With this important gap identified, it calls for the urgent need to examine the perceptions, challenges and opportunities of these community-based education programs for promoting interprofessional education.

### Aim

The study aims to examine the perception, challenges and opportunities of community-based education programs for promoting Interprofessional education in the University for Development Studies.

### Research Questions

#### Main research question

What are the perceptions, challenges, and opportunities of UDS health students and faculty regarding community-based education programs for interprofessional education?

#### Specific research questions

How do health professional students and faculty at UDS perceive the role of community-based education programs in fostering interprofessional education?

What challenges do health professional students and faculty at UDS identify in utilizing community-based education programs as a platform for fostering interprofessional education?

What opportunities for interprofessional education are identified by health professional students and faculty at UDS through their experiences with community-based education programs?”

### Objectives of the study

#### Main Objective

To explore the perceptions, challenges, and opportunities among UDS health professionals and faculty regarding community-based education programs for interprofessional education.

#### Specific Objectives

To explore the perceptions of health professional students and faculty at UDS regarding the role of community-based education programs in fostering interprofessional education

1. 2. To identify the challenges experienced by health professional students and faculty in utilizing community-based education programs as a platform for interprofessional education.
2. To examine the opportunities perceived by health professional students and faculty at UDS for promoting interprofessional education through community-based education programs.

### Significance of the Study

This study will contribute to the role of community-based education programs towards advancing interprofessional education (IPE). Also, examining the contribution of such programs in the development of interprofessional teamwork among students in healthcare, the study will gain insight into the performance of such programs in preparing students for real collaborative practice in healthcare. Besides, the study will identify challenges and opportunities needed to fix interprofessional education within these programs. The findings will inform the refinement of learning activities, which in turn will drive better healthcare delivery through enhanced cooperation and collaboration by health professionals.

## METHODOLOGY

### Research philosophy

Interpretivist paradigm will be the foundation of this research, which supposes reality to be constructed by human interaction, experience, and interpretation (Schwandt, 2000). Unlike objective-truth-seeking positivist paradigms reliant on quantitative variables, interpretivism is concerned with understanding the subjective meanings that individuals make of their experienced life in specific settings (Denzin & Lincoln, 2018).

This philosophical stance is best for studying how health professional students and faculty perceive community-based education programs (COBES and TTFPP) as platforms for promoting Interprofessional Education (IPE). As IPE is inherently a shared and collaborative learning endeavor, the interpretivist stance is a proper foundation for researching how individuals experience and interpret their roles, interactions, and learning in interprofessional learning settings (Berger & Luckmann, 2016).

The focus of the study on contextual understanding and meaning-making is well-aligned with this paradigm, as it allows the researcher to personally immerse within the nuanced points of view, assumptions, and social interactions constituting participants’ experiences. In capturing these in-depth dynamics, the results will reflect participants’ real-life teaching environments and are able to contribute contextually supported recommendations to inform curriculum development, institutional change, and the encouragement of interprofessional practice at UDS and similar institutions (Creswell & Creswell, 2018).

### Research approach

This study will employ a qualitative research approach that is most appropriate for exploring the perceptions, challenges, and opportunities regarding the community-based education programs (COBES and TTFPP) as a platform for promoting interprofessional education (IPE). Qualitative research is focused on making sense of human experiences by attempting depth at the cost of quantification in terms of numbers (Creswell & Creswell, 2018). With IPE involving collaborative learning and social exchanges, a qualitative approach will enable the researcher to capture rich, context-dependent realities of UDS health professionals students and faculty members (Denzin & Lincoln, 2018).

This will facilitate intense scrutiny of participants’ lived experiences, whereby the extent to which community-based education programs facilitates or hinders interprofessional collaboration can be more appropriately understood (Bryman, 2012). While quantitative research seeks to test hypothesis, qualitative research is more suited to uncover emergent findings through firsthand interaction with participants (Charmaz, 2014). With the use of this approach, the study will yield rich, detailed data that will inform recommendations on how to improve the integration of IPE into community-based education programs at UDS and across many health training institutions in the country (Saunders et al., 2016).

### Research design

This study will employ a phenomenological research approach to explore UDS health professional students’ and faculty members lived experiences regarding the community-based education programs (COBES and TTFPP) as platforms for interprofessional education (IPE). This study design is appropriate because it focuses on how individuals interpret their experiences, thus well placed to unveil the complexities of interdisciplinary learning (Creswell, 2013).IPE emphasizes collaboration and hence, phenomenology study design will allow the researcher to investigate participants’ experiences, challenges, and opportunities in community-based education programs (COBES and TTFPP) for promoting IPE. Phenomenology provides rich, qualitative descriptions that cannot be obtained using quantitative studies, and thus a deeper understanding of the development of teamwork and interprofessional learning in actual settings (Padilla-Diaz, 2015).

### Study area/Setting

The study will be done at the University for Development Studies (UDS) Tamale Campus, which is situated in the Dungu suburb of the Tamale South District of the Northern Region of Ghana. The campus’s geographical location is 9.3723° N and 0.8852° W, and its GPS address is NT-0272-1946. UDS is a respected public university in Ghana, mandated by PNDC Law 279 of 1992, and is highly recognized for complementing academic education with societal service through its unique field-based education approach. The Tamale Campus has some of the health-related faculties, including the School of Medicine, School of Allied Health Sciences, School of Pharmacy, and the School of Nursing and Midwifery. These faculties provide undergraduate and postgraduate studies such as Medicine, Nursing, Midwifery, Nutrition, and Pharmacy. Two broad community-based education programs underlie this study: The Third Trimester Field Practical Programme (TTFPP) and the Community-Based Education and Service (COBES) program. TTFPP is a university-wide initiative that positions students from a variety of academic backgrounds including health, agriculture, education, and development studies into communities each year to engage in experiential, interdisciplinary problem-solving. Though it begins as an interprofessional experience, it becomes increasingly discipline oriented as the years go by. Community-based education and services (COBES), on the other hand, is a community-based educational approach followed only for medical students to expose them to rural healthcare problems and encourage service learning. Despite its alignment with the ethos of interprofessional education (IPE), COBES has not been replicated with other health professional students. UDS, whose long history has been community-focused training and diversified portfolio of health programs, has a good solid and proper backdrop for this investigation. To have both TTFPP and COBES represents a rare position to study under which circumstances interprofessional learning may be embedded in and scaled across existing systems. The research will be directed at the health schools including; medicine, allied Health Sciences, pharmacy, nursing and midwifery schools making UDS the optimal location for identifying the perceptions, challenges, and prospects for improving IPE through the community-based education programs (COBES and TTFPP).

### Population of the study

The study sample will consist of final-year health professional students and faculty from the University for Development Studies (UDS). Final-year students who are enrolled in medicine, nursing, pharmacy, nutrition, and midwifery programs, and who have taken part in either the Community-Based Education and Service (COBES) or the Third Trimester Field Practical Programme (TTFPP), will be eligible for sampling. Final-year students are approached because they have been exposed to the full cycle of UDS’s community-based training programs and thus are best placed to critically reflect on their experiences. Their maturity and acquired theoretical and practical knowledge, and broader perspective of interdisciplinary learning environments place them in the best position to investigate perceptions of interprofessional education. Faculty members teaching and mentoring students enrolled in these programs will also be included, as their viewpoints are crucial to learning about community-based education programs as platforms for advancing interprofessional education.

### Inclusion criteria

The final -year medical students who have completed the COBES and TTFPP courses will be recruited to the study. Final-year nursing, nutrition, pharmacy, and midwifery students who have completed the TTFPP course will also be selected. Only those who provide informed consent will be included.

Members of the faculty who have taught and mentored the TTFPP program from all fields of health, including medicine, nursing, pharmacy, nutrition, and midwifery, will be the study participants. This will include faculty with direct roles in COBES, particularly the medical school, whose rich experiences and expertise will be infused. Faculty members from other health programs will also provide their understanding of community-based education for improving interprofessional education (IPE).

This sampling will give a broad and representative array of perspectives, capturing data from students and faculty members who are directly involved in these community-based education programs. Having faculty members from various health disciplines will add richness to the findings and give a balanced picture of the role of COBES and TTFPP towards interprofessional education.

### Exclusion criteria

Health professionals students and faculty members who may be ill or have physical or mental health issues while collecting data will be excluded. Students and faculty who may not be present for data collection throughout the study duration.

### Variables of the study

Third Trimester Field Practical Programme (TTFPP) and Community-Based Education and Service (COBES) perceptions, knowledge of Interprofessional Education (IPE), TTFPP and COBES experiences, opportunities for interprofessional learning, challenges in the programs, and recommendations for enhancing IPE through TTFPP and COBES will be the central variables of interest for this study.

### Sample Size

The sample size of this study will be guided by the data saturation principle, the point where no further themes or insight can be elicited from the data (Guest, Bunce, & Johnson, 2006). In order to get a good range of opinions regarding the perception of the community-based education programs (COBES and TTFPP) as platform for promoting IPE, approximately ten faculty members representing a diverse range of health professions will be selected to participate in the study. In addition, an estimated twenty final-year health professionals students from the various health programs including pharmacy, nutrition, midwifery, nursing, and medicine will be chosen on a purposeful basis. This diversity of student samples is intended to enhance response variability and represent a wide range of experiences (Saunders, Julius, & Tom, 2017). According to previous qualitative research, which argues that a sample size of fifteen to thirty participants is typically best for phenomenological studies, the proposed sample is considered adequate for producing rich and meaningful data (Marshall, Cardon, Poddar, & Fontenot, 2013).

### Sampling procedure

This study will employ purposive sampling, a non-probability sampling technique to select respondents with expert knowledge or experience pertinent to the research (Patton, 2015; Etikan et al., 2016). Purposive sampling is particularly suitable for this study because it allows respondents with direct experiences of community-based education programs (COBES and TTFPP) to be selected in a manner that ensures data collected are rich and meaningful to the research objectives. For purposes of ensuring sample representation, the study will recruit participants from various health professions (medicine, nursing, pharmacy, nutrition, and midwifery) as well as different faculty members with varying experience levels in supervising community-based education (COBES and TTFPP). This will ensure that results are generalizable to the greater population of UDS’s health professional students and faculty to aid the study’s findings to be generalized to such contexts (Marshall et al., 2013).

### Data collection procedures

Permission will be obtained from the university and departments before the collection of the data. Departmental coordinators will assist in identifying potential participants who will be contacted and made aware in full of the purpose, procedures, and ethical considerations in the study. Informed consent will be obtained from all participants before the interviews, ensuring that they are informed of their rights, including withdrawing at any time without penalty. One-to-one in-depth interviews will be conducted in a comfortable and private setting, a private room on campus, to ensure confidentiality and minimize distraction. Each interview will take approximately 30 to 45 minutes to enable participants to give sufficient information about their experiences, perceptions, and concerns of these community-based education programs (COBES and TTFPP). The interviews will be conducted with the aid of an interview guide and all sessions will be audio-recorded with the consent of the participants to ensure accuracy and completeness in data gathering. Apart from audio recordings, the field notes will be recorded by the researcher to also pick up on non-verbal cues, context, and notes that may be useful in aiding further understanding of participants’ responses.

Data collected will be stored securely in password-protected files, and personal details will be removed to ensure confidentiality. Audio recordings will be transcribed verbatim for analysis, and the transcripts will be compared with the recordings to ensure accuracy. This systematic process in gathering the data will render the findings reliable and valid (Creswell, 2018).

### Data collection tools/instruments

This study will use face-to-face in-depth interviews as the primary data collection tool. The use of interviews for this research is appropriate given that it provides a means of getting respondents to speak about personal experiences, observations, and perceptions about these community-based education programs(COBES and TTFPP) in their own terms, producing rich contextual information (Creswell, 2018).

An interview guide will be developed to ensure consistency in questioning but with the provision for flexibility for follow-up discussion. For guiding the research instrument design, literature review was conducted prior to constructing the interview guide. Variables employed under research objectives were used as a reference for the questions. The interview guide will be in the form of open-ended questions to obtain participants’ perceptions, challenges, and opportunities for community-based education programs (COBES and TTFPP) as a platform for interprofessional education.

### Pre-testing/Pilot testing

To ensure the validity and reliability of the study findings, some health professional students and faculty members with characteristics similar to the targeted participants will be utilized for pre-testing the interview guide but will be excluded from the main study. Since they have been pre-exposed to the interview questions already, pre-test participants might react differently to actual data collection and therefore their participation in the main research might lead to bias.

Excluding them in the primary data will produce current, unbiased findings and representative of the larger study population. To increase the quality, relevance, and readability of the questions, pre-test feedback will be carefully examined and included. This will improve the overall quality and effectiveness of the data collection tool.

### Data analysis

Analysis of the data in this study will be conducted in relation to thematic analysis with Nvivo software, which is applicable for qualitative studies (Braun & Clarke, 2022). Thematic analysis can allow an individual to code, investigate, and report on qualitative data patterns (themes). Thus, the results can capture best participants’ perceptions and experiences.

Data analysis will encompass six general steps: familiarization with data, initial coding, theme searching, exploration of themes, naming and labeling themes, and final report writing (Braun & Clarke, 2022; Creswell, 2018). The researcher will carry out verbatim interviews transcription and read repeatedly to build an in-depth comprehension of data. Data coding will also be done through Nvivo computer software to allow for effective storage and retrieval of data to support extensive and systematic thematic analysis (Miles & Huberman, 1994).

To achieve validity and reliability, peer debriefing and member checking will be applied. Peer debriefing involves sharing emerging themes with peers and supervisors in order to make interpretations (Enworo, O. C. (2023). Member checking will allow participants to review findings summaries to make sure that interpretations were accurate (Creswell, 2018).

Triangulation of data will also be done through cross-matching results in different sources of data, one-on-one interviews such that the quality of findings would be determined (Patton, 2015). Final analysis will provide a thorough, descriptive account of participants’ experience such that the study findings will be valid as well as transferable to environments alike.

### Sources of data

In the interests of this study, only the primary data will be employed for the anal. It will be sourced from first-hand research respondents through the application of in-depth interviews (Denzin & Lincoln, 2011). These practices will facilitate collection of firsthand experience and data used during research regarding the community-based education programs (COBES and TTFPP) as an additional platform for interprofessional education (IPE). The responses of the participants will offer a collective understanding of the interdisciplinary collaboration that exists during the community-based education programs. Through the utilization of only the primary data, the study will ensure the validity and relevance of the findings, resulting in a better understanding of the research questions.

### Ethical considerations

Ethical clearance will be issued by the University for Development Studies Institutional Review Board (IRB) before collecting data.

The participants will be given a clear explanation of the reason for the study, and consent will be requested before they are interviewed. They will be participating voluntarily and will be able to withdraw at any time without penalty. To ensure confidentiality, the identity of the participants will not be disclosed, and all information will be kept securely with restricted access.

### Study Status and Timeline

No participants have been enrolled and no data gathering has been initiated as of submission. Ethical approval for the research is awaiting review and should be submitted by July 2025. Participant recruitment will be done in August 2025, while data collection is hoped to take place between September and October 2025. Data transcription and analysis will be carried out between October and November 2025, while the study report will be ready by December 2025. Defense and final submission of the thesis will be between January–February 2026. No results have been generated at any stage of the study. This protocol thus meets the PLOS ONE requirements for study protocols.

### Trustworthiness of the Study

For assuring trustworthiness, this study follow earlier qualitative research standards. White (2005) writes that credibility of qualitative research is attained through assuring the research process to be free from bias and confidence regarding findings is assured based on the research design. This study assure trustworthiness through the following primary standard. To provide credibility, member checking will be employed in the research whereby participants check for accuracy through reading summaries of their interviews. Aiming for prolonged engagement with the data will involve repeated readings of transcripts to ensure authenticity. Transferability will be maximized using full descriptions of research context, study participants, and research activities such that other researchers can make their own determination as to the relevance of the results to similar settings. To enhance reliability, the research process will be thoroughly documented, including data collection procedures, transcription, and analysis processes. Peer debriefing with supervisors and colleagues will guarantee consistent interpretation.

Confirmability will be achieved through maintaining an audit trail, documenting every decision that is made within the research process. Data triangulation, in incorporating field notes and interview transcripts, will also facilitate the neutrality and reliability of findings.

### Researcher reflexivity

Placing reflexivity within the context of this study, which explores the possibility of integrating interprofessional education (IPE) within community-based education programs (COBES and TTFPP), the researcher will locate himself as an active member within the research process and the potential for bias within data collection and analysis. In addressing this, the researcher will employ several strategies to ensure reflexivity and maximize the trustworthiness of the research.

The researcher will also maintain a reflective journal during the research. The journal will be utilized to record the researcher’s assumptions, personal reactions, and reflections during the process of conducting interviews, data analysis, and interpretation of findings. It will enable the researcher to maintain an awareness of personal biases by reflecting over the personal preconceptions (Berger, 2015).

Further, the researcher will also go through peer debriefing with peers and supervisors. In these meetings, initial themes and interpretations will be shared and critically evaluated. This collective thinking will help identify and address any surfacing biases to ensure validity and credibility of findings (Creswell, 2018). By these processes, the researcher will guarantee openness and rigor in the research process, which will result in a more credible and trustworthy study.

Timelines and milestone

|  | **Activity** | **Timeline** |
| --- | --- | --- |
| **1** | Submission<br>for IRB<br>Ethical<br>Approval | **June,2025** |
| **2** | IRB<br>Approval<br>Expected | **July,2025** |
| **3** | Development<br>and Pre-testing of<br>Interview<br>Guide | **August** |
| **4** | Participant<br>Recruitment | **August** |
| **5** | Data<br>Collection<br>(Interviews | **Septembe**<br>**r** |
| **6** | Data<br>Transcription<br>and Cleaning | **October** |
|  | Data analysis | **October** |
| **7** | Member<br>Checking<br>and<br>Validation | **November** |
| **8** | Final Report<br>Writing | **November** |
| **9** | Data Upload<br>to<br>Repository | **December** |
| **10** | Thesis<br>submission<br>and defense | **January-**<br>**February** |

### Expected Results

As this is a study protocol and no data has been gathered or analyzed, the subsequent are anticipated findings based on the study objectives and evidence in the current literature.

Objective 1: To explore the perceptions of health professional students and faculty at UDS regarding the role of community-based education programs in fostering interprofessional education.

It is anticipated that participants will share positive views of community-based learning as a foundation for Interprofessional education.

From evidence available in similar international studies, it is expected that both students and faculty at the University for Development Studies (UDS) will have general positive perceptions of the potential of community-based education (CBE) programs, such as COBES and TTFPP, to promote Interprofessional Education (IPE).

Students need to emphasize the interactive and experiential nature of such programs in playing a fundamental role in the development of interprofessional competencies like communication, teamwork, and respect for other professional roles. Suwanchatchatchai et al. (2024), for instance, reported notable change in communication, conflict resolution, and team working in students following exposure to a structured Community-Based Interprofessional Education (CBIPE) module. Similarly, Eggenberger et al. (2019) demonstrated that community-based interdisciplinary projects enabled professionals to better understand and appreciate diversity.

Faculty perceptions are also expected to legitimize the educational value of CBE in promoting integrated, patient-centered care and collaborative attitudes among health professions students. Some issues can, nonetheless, be raised about the lack of preparation for interprofessional facilitation, institutional constraints, and limited consistent administrative support (Kristina et al., 2023; Eggenberger et al., 2019).

Objective 2: To identify the challenges experienced by health professional students and faculty in utilizing community-based education programs as a platform for interprofessional education.

It expects to encounter structural, logistical, and pedagogical issues that are usually caused by interprofessional learning in community environments.

The study is anticipated to reveal problems such as fragmented program structures, out-of-sync department calendars, and insufficient time for interprofessional collaborative learning. Such problems tend to impact the development of long-lasting and meaningful interprofessional teams (Eggenberger et al., 2019; Suwanchatchai et al., 2024). Students also tend to experience problems related to role confusion, uneven levels of participation, and lack of support in team learning.

Faculty problems may include insufficient exposure to interprofessional pedagogical strategies and insufficient coordinated professional education. As discovered in related literature, scholars are inclined to stick to conventional teaching models that may be weakly integrated with collaborative learning contexts (Suwanchatchai et al., 2024).

The study can also pinpoint logistical constraints such as scarce resources, restricted vehicle access, and inadequate supervisory coverage. These issues have a tendency to limit reach and sustainability of CBE-IPE programs. Keshmiri and Barghi (2021) underscored that infrastructure gaps and binary funding of programs can limit complete integration of interprofessional methods. Further, social and institutional hierarchies, particularly those favoring some professions over others, can reduce inter-professional interaction and affect team dynamics (Randita et al., 2019).

Short-term local community placements, as intensive as they may be, are likely to restrict the evolution of trust and familiarity with teams. Absence of formal procedures for evaluating interprofessional practice, like absence of standardized evaluation tools, can also hinder feedback cycles and improvement planning.

Objective 3: To examine the opportunities perceived by health professional students and faculty at UDS for promoting interprofessional education through community-based education programs.

The study anticipates participants to suggest practical, institutional, and technological means of ensuring interprofessional collaboration through existing programs.

The outcomes are expected to outline some options for building interprofessional learning by taking advantage of existing community-based programs. The participants may suggest incorporating combined service projects involving contributions from multiple disciplines, especially in tackling multifaceted health issues at the community level. Suwanchatchai et al. (2024) and Kristina et al. (2023) demonstrated how collective accountability within health interventions enabled enhanced team participation and prolonged collective learning.

Students and facultyalso see opportunities for increasing interdisciplinarity communication through reflective debriefing, peer mentoring, and community workshops. Eggenberger et al. (2019) reported that such collaborative processes promoted more standardized interaction and understanding of different roles in health.

Expanded digital spaces can facilitate inter-disciplinary teams to communicate continuously, particularly where face-to-face collaboration is impracticable. The technology can facilitate real-time planning, case discussion, and collective assessment.

Administrative coordination and faculty development are similarly apt to enter into focus as achievable opportunities. Creating institutional structures that promote cross-disciplinary supervision, models of co-teaching, and interprofessional performance evaluation is able to foster the culture of collaboration (Keshmiri & Barghi, 2021).

The COBES and TTFPP models, as they are practiced today in UDS, lay a base for the building of more robust IPE. Strategic alignment in cross-disciplinary integration can potentially enable more integrated and contextualized training for the next generation of health workers.

Stakeholders might champion reimagined placements, shared learning goals, and interprofessional field teams to improve health outcomes and team readiness at graduation.

### Summary

The expected outcomes emphasize UDS community-based education programs’ potential for supporting health professions students’ and faculty members’ development of interprofessional competencies. Both groups’ views are predicted to demonstrate a strong value placed on experiential and community-focused learning environments promoting professional interaction, teamwork, and context-specific learning.

Several structural and systemic problems can be determined, including disconnected curricula, siloed institutions, faculty preparedness, and logistical limitations. These challenges, though persistent, can be addressed by intentional program redesign, alignment among stakeholders, and interprofessional training capacity funding.

Participants will also be able to recognize a range of opportunities to more fully integrate IPE into existing community-based systems. These include interdisciplinary projects, reflective practices, technology-enabled collaboration, and faculty development. Taken together, the opportunities identify the potential for strengthening interprofessional learning within the UDS environment and furthering the institution’s contribution to educating health professionals for interprofessional collaborative practice in diverse and underserved settings.

## Discussion

Specially crafted community-based education programs infusing interprofessional education principles have the potential to significantly improve the learning trajectories of health professions students. Within the context of the University for Development Studies (UDS), field-based programs such as the Community-Based Education and Service (COBES) and the Third Trimester Field Practical Programme (TTFPP) provide experiential learning opportunities that are rich in nature. These programs are intended to allow students to appreciate the practice of interdisciplinarity, with on-the-job training in interprofessional collaboration to address community health challenges. The interactivity of such field placements is likely to engender more professional understanding of their roles, enhance communication, and promote mutual respect among interprofessional students (Suwanchatchai et al., 2024). The faculty themselves will probably appreciate these benefits, as well, particularly with respect to developing teamwork thinking and practice-readiness in students. Nevertheless, the absence of formalized interprofessional learning objectives and set mechanisms for collaboration will temper the full realization of these benefits. Institutional culture, including supervision norms, curriculum teaching, and reflective practice, is significant in influencing the effectiveness of such community-based programs toward the achievement of interprofessional learning outcomes (Eggenberger et al., 2019).

The integration of interprofessional education into community-based learning environments is not without challenges. Inconsistency between curricular designs and academic schedules among departments can complicate the planning of interprofessional field experience (Keshmiri & Barghi, 2021). Students could end up having disconnected learning experiences in which opportunities to engage with students from other disciplines are ambiguous or superficial. Staff, although supportive in concept, may be unskilled in pedagogy to facilitate interprofessional practice, which is bound to require deviation from evidence-based discipline-specific education (Eggenberger et al., 2019). Moreover, budgetary constraints involving mobility problems, inadequate supervision, and inadequate economic support for extended placements can reduce the intensity and duration of these programs. Hierarchical norms in health professions that are inclined to support the functions of particular disciplines can also inhibit equitable participation and collaborative decision-making processes when in practice (Randita et al., 2019). The obstacles are then compounded by the absence of standardized measures for quantifying interprofessional competencies so that it is difficult to track students’ progress or program impact. Hence, what is required is a comprehensive institutional commitment, one that involves interdepartmental collaboration, staff development, operational assistance, and curriculum redesign aimed at fostering sustainable interprofessional learning environments.

In spite of these limitations, there are several fruitful ways interprofessional education can be enhanced in the context of UDS’s community-based programs. Field placements can be reorganized to include purposefully designed interprofessional projects, where students of different disciplines work together to conduct health promotion, community diagnosis, or service delivery tasks (Suwanchatchai et al., 2024). These projects would not only enrich students’ learning experiences but also address authentic community health priorities through interprofessional, team-based approaches. The inclusion of structured debriefing sessions, peer mentoring, and interdisciplinary reflection in fieldwork can also further consolidate interprofessional competencies. Technology also provides a state-of-the-art platform for enhancing interprofessional collaboration, especially for geographically dispersed or resource-constrained settings. Virtual simulations, teleconsultations, and collaborative planning tools can potentially provide continuity of student engagement and facilitate cross-disciplinary discussion (Eggenberger et al., 2019). Developing faculty skills through targeted training in interprofessional facilitation, evaluation strategies, and interprofessional pedagogy is equally important. This can involve co-teaching models, shared supervision frameworks, and interprofessional curriculum boards. Leaning on the robust infrastructure and long-established tradition of experiential education at UDS further provides COBES and TTFPP with a great platform to be an example for interprofessional training across sub-Saharan Africa. With careful redesign and more stakeholder involvement, these programs could be the foundation for building a cooperative, community-oriented health workforce that will solve the complex healthcare problems.

## Conclusion

This protocol aims to find out how University for Development Studies community-based education programs can be used to provide interprofessional education to students of the health professions. By comparing and contrasting the perceptions of different students and faculty members, the study aims to find out existing perceptions, challenges, and opportunities in the COBES and TTFPP environments. The expected results show a great potential for the programs to foster interprofessional collaboration, provided that specific institutional and pedagogical challenges are resolved. Areas of priority in program development are revealed in the discussion, including harmonization of curriculum design, faculty development, logistical coordination, and introduction of formal interprofessional learning activities. In recreating those community-based programs through an interprofessional lens, UDS can foster a new model of health education that prepares future practitioners for collaborative and socially accountable service provision.

## Data Availability

No datasets were generated or analysed during the current study. This manuscript presents a qualitative research protocol and does not report results or pilot data at this stage. All relevant data from this study will be made available upon completion of the research. Following data collection and analysis, deidentified transcripts and thematic summaries will be stored in a secure institutional repository at the University for Development Studies (UDS), Ghana. Access to these materials will be provided in accordance with ethical guidelines and participant confidentiality agreements. Upon publication of the full study results, deidentified data—such as anonymized interview transcripts, coding frameworks, and supporting qualitative datasets—will be made publicly available through the UDS Research Repository or other recognized open-access repositories such as Zenodo or Figshare. The repository link, DOI, and access instructions will be updated in the final published article and supporting information. Any researchers seeking access to confidential data prior to public release may direct their request to the Institutional Review Board (IRB) of the University for Development Studies at. Access may be granted to researchers who meet the criteria for access to sensitive data, including having ethical approval from a recognized research body and a commitment to protect participant anonymity. All data shared will comply with the requirements of the UDS-IRB, and no identifiable personal information will be disclosed. Data will be retained and preserved in accordance with UDS institutional policy and Ghanaian national data protection regulations.

## Declarations

### Ethical approval and consent

Ethical clearance for this study will be obtained from the Institutional Review Board (IRB) of the University for Development Studies. All participants will be required to give informed consent before participating in the study. Confidentiality and anonymity will be ensured, and participants will be made aware of their right to withdraw from the study at any point without penalty.

### Author contributions

MAA, BAA, and PAAK conceptualized and designed the study. MAA prepared the first draft of the protocol.VM, and SDZ provided critical revisions and intellectual input. All authors reviewed and approved the final manuscript.

## Acknowledgments

The research team acknowledges the support of the Department of Health Professions Education and Innovative Learning at UDS, as well as the faculty and students who will be participating in this study.

